# *In-silico* modeling of COVID-19 ARDS: pathophysiological insights and potential management implications

**DOI:** 10.1101/2020.07.21.20158659

**Authors:** Anup Das, Sina Saffaran, Marc Chikhani, Timothy E Scott, Marianna Laviola, Nadir Yehya, John G. Laffey, Jonathan G. Hardman, Declan G. Bates

## Abstract

**Objectives:** Patients with COVID-19 Acute Respiratory Distress Syndrome (CARDS) appear to present with at least two distinct phenotypes: severe hypoxemia with relatively well-preserved lung compliance and lung gas volumes (Type 1) and a more conventional ARDS phenotype displaying the typical characteristics of the ‘baby lung’ (Type 2). We aimed to test plausible hypotheses regarding the pathophysiological mechanisms underlying CARDS, and to evaluate the resulting implications for ventilatory management.

**Design:** We adapted a high-fidelity computational simulator, previously validated in several studies of ARDS, to (a) develop quantitative insights into the key pathophysiologic differences between CARDS and conventional ARDS, and (b) assess the impact of different PEEP, FiO_2_ and tidal volume settings.

**Setting:** Interdisciplinary Collaboration in Systems Medicine Research Network.

**Subjects:** The simulator was calibrated to represent CARDS patients with both normal and elevated body mass indices undergoing invasive mechanical ventilation.

**Measurements and Main Results:** An ARDS model implementing disruption of hypoxic pulmonary vasoconstriction and vasodilation leading to hyperperfusion of collapsed lung regions failed to replicate clinical data on Type 1 CARDS patients. Adding mechanisms to reflect disruption of alveolar gas-exchange due to the effects of pneumonitis, and heightened vascular resistance due to the emergence of microthrombi, produced levels of V/Q mismatch and hypoxemia consistent with data from Type 1 CARDS patients, while preserving close to normal lung compliance and gas volumes. Atypical responses to PEEP increments between 5 and 15 cmH_2_O were observed for this Type 1 CARDS model across a range of measures: increasing PEEP resulted in reduced lung compliance and no improvement in oxygenation, while Mechanical Power, Driving Pressure and Plateau Pressure all increased. FiO_2_ settings based on ARDSnet protocols at different PEEP levels were insufficient to achieve adequate oxygenation. Incrementing tidal volumes from 5 to 10 ml/kg produced similar increases in multiple indicators of ventilator induced lung injury in the Type 1 CARDS model to those seen in a conventional ARDS model.

**Conclusions:** Our model suggests that use of standard PEEP/ FiO_2_ tables, higher PEEP strategies, and higher tidal volumes, may all be potentially deleterious in Type 1 CARDS patients, and that a highly personalized approach to treatment is advisable.

## Introduction

COVID-19 has challenged the global community with a novel disease that is currently not fully understood. As well as the elderly, it has a predilection for the co-morbid, particular ethnicities, and the obese. COVID-19 ARDS (CARDS) appears to have a dynamic, time-related, disease spectrum with at least two proposed ‘phenotypes’: a Type 1 (non-typical ARDS), characterized by near-normal lung compliance and pulmonary gas volume, combined with large shunt fraction and severe hypoxemia [1,2,3], which may evolve into, or exist alongside, a Type 2 phenotype characterized by a lower lung compliance (‘baby lung’ [4]) and a general clinical presentation more typical of severe ARDS. Pulmonary microvascular thrombosis and associated ischemic events also appear to be a particular characteristic of the disease [3].

It is currently unclear whether transitions from the Type 1 to Type 2 phenotype are due to the natural evolution of the disease, or to damage to the lung resulting from inappropriate initial ventilatory management leading to a cycle of progressively injurious ventilator-induced lung injury (‘VILI vortex’) [5,6], or to a combination of the two. Wide variations in mortality rates across different intensive care units, together with unexpected patient responses to usual guidelines for ventilator management, have led some clinicians to assert that standard ventilation protocols employing PEEP/FiO_2_ tables, ‘open lung’ targets, etc., may need significant revision, and selective application of their components, in CARDS patients [6]. A primary concern is that pathophysiological changes underlying ‘classical’ ARDS ventilation approaches may not apply in some CARDS patients, and may thereby lead to ventilator induced injury, as represented by the conversion of patients from the Type 1 to Type 2 phenotypes. For example, the use of relatively high PEEP in ARDS treatment is based on the assumption that hypoxemia is due to recruitable, non-aerated lung regions; however, in the Type 1 CARDS patient, the lung units may be open but non-functional due to vascular shunt, and thus PEEP may over-distend (rather than recruit) these lung units, leading to injury without any improvement in gas exchange. The avoidance of VILI in CARDS patients is also of particular importance given the extended time periods for which many of these patients receive mechanical ventilation.

Based on insights gained from clinical experience, hypotheses have been put forward to explain the particular pathophysiological characteristics of CARDS patients. In [1,2], pulmonary vasoparesis (i.e. globally reduced pulmonary vascular tone) resulting from impaired regulation of lung perfusion and disruption of hypoxic pulmonary vasoconstriction (HPV) was proposed as a possible explanation for the severe hypoxemia occurring in the compliant lungs of Type 1 patients, while results in [18-20] indicate a potentially important role for endothelial damage due to pneumonitis and microthrombi. However, there is currently no direct evidence with which to quantitatively evaluate the effect on gas-exchange and V/Q mismatch of each of these mechanisms, alone or in combination. The clinical evaluation of alternative ventilation strategies for CARDS based on hypothesized models of its pathophysiology is also extremely difficult in the current circumstances.

To shed light on the above issues, we employed a multi-compartmental computational model that simulates highly integrated pulmonary and cardiovascular physiology together with a detailed representation of the effects of invasive mechanical ventilation. This model has been successfully deployed in several previous studies investigating the pathophysiology and ventilatory management of conventional ARDS, [7-13]. In this study, we use this model to simulate proposed pathophysiological mechanisms underlying CARDS, by altering key parameters to create exemplar patients that reproduce the currently available clinical data. The biomechanics and physiology of obesity are readily amenable to study within this model, and so we include an exemplar patient from this at-risk group. We then use these models to compare the effects of different ventilator settings for CARDS and conventional ARDS patients.

## Methods

### Cardiopulmonary computational model

The study employs a multi-compartmental computational model that simulates a dynamic *in vivo* cardiovascular-pulmonary state, comprising conducting airways and a respiratory zone of 100 parallel alveolar compartments, with each compartment having an independent set of parameters accounting for stiffness, threshold opening pressures (TOP) and extrinsic pressures that compress the alveolus as well as airway and peri-alveolar vascular resistances. This allows for a wide spectrum of ventilation perfusion mismatch to be replicated. The model includes inherent physiological reflex mechanisms, e.g. hypoxic pulmonary vasoconstriction. The capability of the simulator to represent the cardiopulmonary disease states of individual patients with ARDS has been validated in a number of previous studies [7-13], and the mathematical and physiological principles on which it is based are detailed in the supplementary material.

### Measurements of key physiological variables and VILI indices from the model

Mechanical ventilation in the model is set to pressure-controlled mode with respiratory rate at 20 bpm, inspiratory to expiratory ratio at 1:2 and FiO_2_ set to 100%. To observe the cardiopulmonary effects of interest, the following values were computed and recorded: arterial oxygen tension (PaO_2_), arterial carbon dioxide tension (PaCO_2_), arterial pH (pH), arterial and mixed venous oxygen saturation (SaO_2_ and SvO_2_), volume and pressure of individual alveolar compartments at end-inspiration and end-expiration (V_alv_insp_, V_alv_exp_, and P_alv_insp_ and P_alv_exp_, respectively), cardiac output (CO), mean arterial and pulmonary artery pressure (MAP and MPAP, respectively), and arterial oxygen delivery (DO_2_; computed using the values of CO, SaO_2_ and PaO_2_ and hemoglobin level (Hb)). Alveolar recruitment was calculated as the fraction of alveoli receiving zero ventilation and subsequently achieving ventilation. VILI indices calculated include respiratory system compliance (C_RS_), intra-tidal recruitment, mechanical power [13], driving pressure, mean alveolar pressure (P_alv_), and dynamic strain (calculated as ΔV / V_frc_, where V_frc_ is V_alv_exp_ at PEEP = 0 cmH_2_O and ΔV = V_alv_insp_-V_alv_exp_).

Intra-tidal recruitment was calculated as the difference between the ratio of total ventilated lung at the end of inhalation to that at the end of exhalation. Shunt fraction (Q_S_/Q_T_) was calculated using the shunt equation, based on the arterial, pulmonary end-capillary and mixed venous O_2_ content. Physiological deadspace (and deadspace fraction (VD/VT)) were calculated from PaCO_2_, mixed expired CO_2_ tension (PE’CO2) and exhaled tidal volume [14]. PEEP_TOT_ is measured as the total pressure within the lung at the end of expiration and accounts for both the extrinsic applied expiratory pressure and intrinsically generated end expiratory pressures. Plateau pressure (Pplat) is measured as the average end-inspiratory pressure in the lung. Lung compliance is calculated as ΔV/ ΔP, where ΔP is the plateau pressure minus PEEP_TOT_. All simulations were run for 30 minutes and during simulations all parameters were updated and recorded every 10 milliseconds.

## Results

### Pathophysiological mechanisms that produce a Type 1 CARDS phenotype

We configured the simulator’s baseline settings to represent two ‘exemplar’ patients – a typical (ideal body weight) 70 kg patient and an obese patient weighing 110 kg (body mass indices 24 kg/m^2^ and 37.7 kg/m^2^, respectively). Patients were assumed to be sedated and to be receiving positive pressure mechanical ventilation without any spontaneous ventilatory effort.

The exemplar patient models were then configured according to clinical data on different CARDS phenotypes available in the literature. Based on the data [1-6] suggesting that Type 1 CARDS patients have relatively well preserved lung gas volume and compliance, the Type 1 models were set to have 8% (11% in patients with Elevated BMI) of their alveoli collapsed, i.e. non-aerated, with low recruitability (i.e. extrinsic pressures acting on collapsed alveolar compartments were set to >40 cmH_2_O with average threshold opening pressures of 50 cmH_2_O), [1-4]. To simulate the hyperperfusion of gasless tissue reported in [1], we implemented vasodilation in the collapsed units by decreasing their vascular resistance by 80%. HPV is normally incorporated in our simulator via a mathematical function, based on the stimulus response curve suggested in [15]; to simulate the hypothesised disruption of HPV in CARDS we disabled this function in our model. Simulating the effects of these mechanisms alone did not produce levels of Q_S_/Q_T_, deadspace and hypoxemia matching those reported in clinical data describing Type 1 CARDS; see Table 1. We therefore also incorporated disruption of alveolar gas-exchange due to the effects of pneumonitis into the model by blocking alveolar-capillary gas equilibration in 30% of the alveolar compartments. As thrombotic complications have been reported to be a characteristic feature of CARDS, we modeled the presence of microthrombi by increasing vascular resistance by a factor of 100 in 10 of the 30 compartments with disrupted gas exchange. Implementing the above additional pathophysiological mechanisms in our model produced levels of Q_S_/Q_T_ (47.8%), deadspace (192.3 ml) and hypoxemia (SaO_2_ 87.6%, FiO_2_ of 100%) that match those reported in clinical data describing Type 1 CARDS [1-6], while still preserving near-normal levels of lung compliance (43.5 ml/cm H_2_O); see Table 1.

**Table 1:**
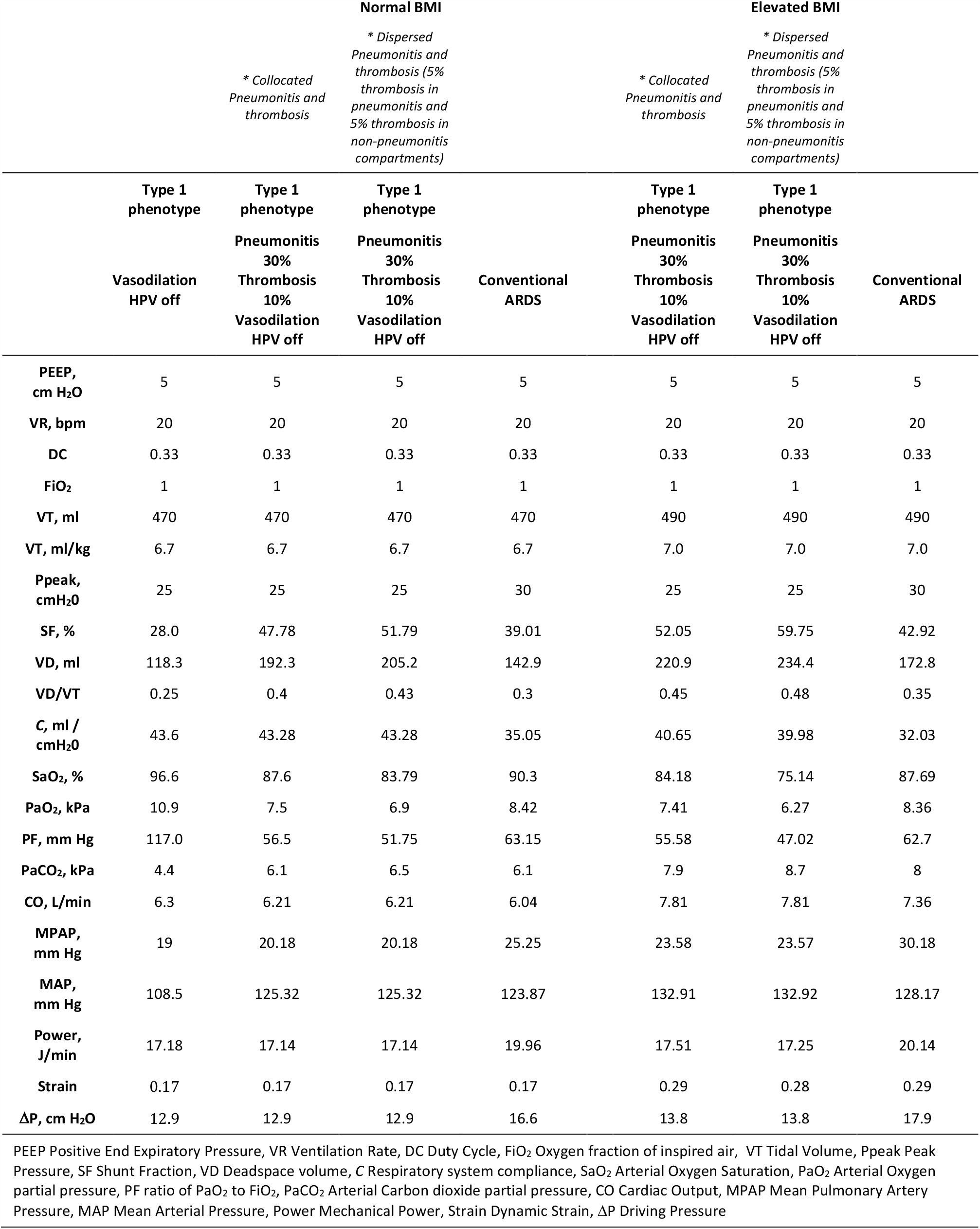
Pathophysiological mechanisms generating a Type 1 CARDS phenotype.

Although a reasonable assumption, there is currently no clear evidence that pulmonary vascular microthrombi primarily arise in alveoli that have been damaged by pneumonitis, and therefore we also modeled the effect of microthrombi being heterogeneously distributed throughout the lung, by increasing vascular resistance in 5 of the disrupted and 5 of the normally functioning compartments. This further worsened V/Q matching (Q_S_/Q_T_ increased to 51.8% and deadspace to 205.9 ml) leading to severe hypoxemia, (SaO_2_ 83.8%, FiO_2_ of 100%); see Table 1.

For the purposes of comparison, both normal and elevated BMI patients were also configured to represent a conventional ARDS phenotype, [3,4]. Specifically, the compliance and threshold opening pressures of 50 alveolar compartments were configured to create non-aerated (at baseline) but recruitable lung regions (extrinsic pressures that induce collapse, set with mean value of 20 cm H_2_O and mean TOP of 20 cm H_2_O), resulting in a Qs/Qt of 39%, VD/VT of 0.3 and compliance of 35.1 ml/cmH_2_O in the normal BMI model.

### Effects of high versus low PEEP

Figure 1 shows the results of reducing PEEP to 5 cmH_2_O, and increasing PEEP to 15 cmH_2_O (from a baseline setting of 10 cmH_2_O), while keeping tidal volume fixed at 7 ml/kg, for both the Type 1 CARDS and conventional ARDS models (normal BMI’s). As shown, strikingly different responses were observed for the two models across a range of measures. For the conventional ARDS model, increasing PEEP produced improvements in oxygen delivery (DO_2_) and arterial oxygenation (PaO_2_ and SaO_2_) resulting from increased alveolar recruitment, increased lung compliance and reduced shunt, with a reduction in driving pressure and only a modest increase in mechanical power. In contrast, for the Type 1 CARDS model, increasing PEEP reduced oxygen delivery, produced no improvement in oxygenation, and reduced lung compliance, while mechanical power, driving pressure and plateau pressure were all increased. In the obese Type 1 CARDS patient, increasing PEEP did slightly increase oxygen delivery and arterial oxygenation, but to a lesser extent than in the conventional ARDS model; see Figure S1 in the Supplementary Material. Corresponding results in terms of relative changes, rather than absolute values, for both figures are shown in Figures S2 and S3 of the Supplementary Material.

**Figure 1:**
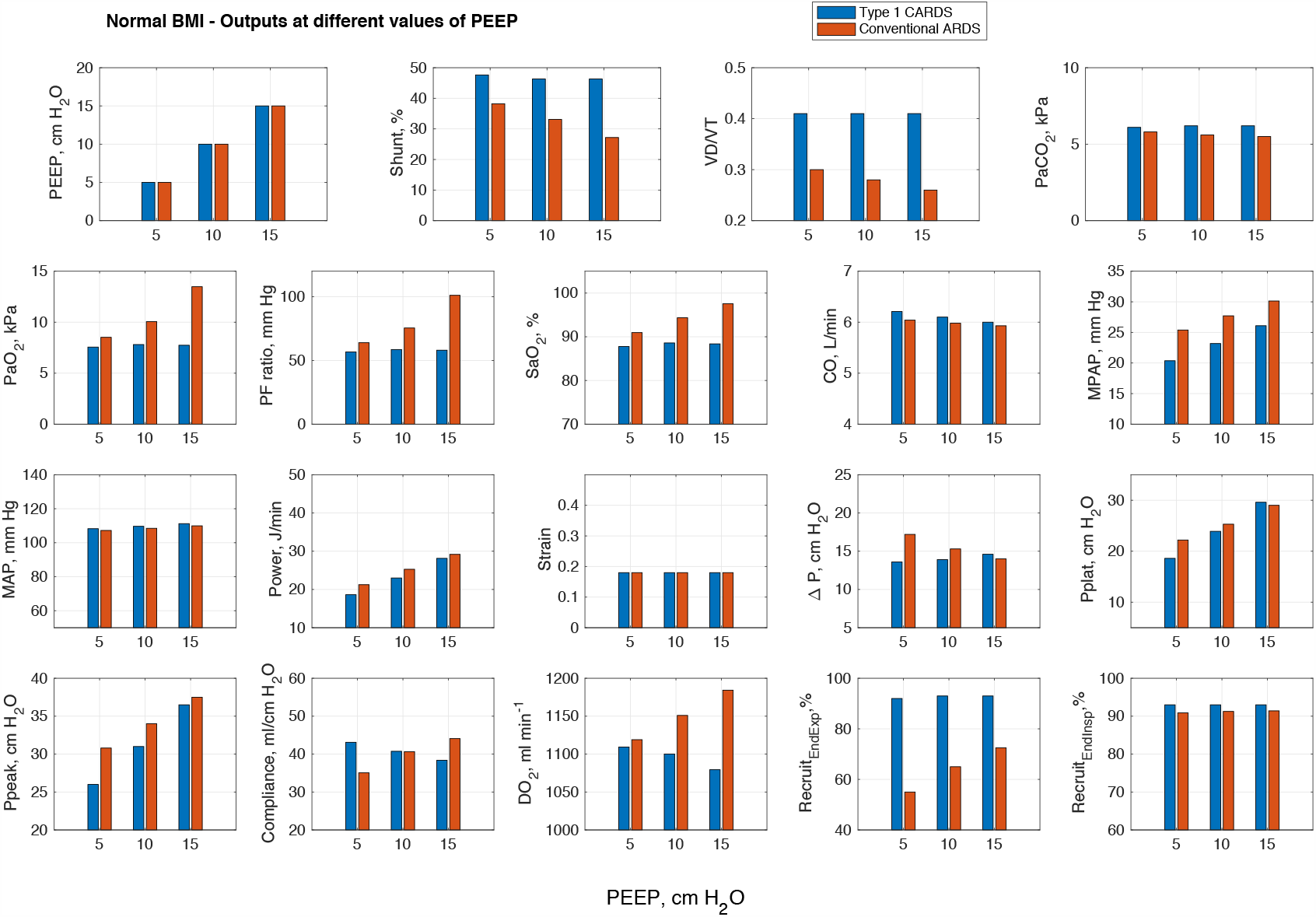
Results of reducing PEEP to 5 cmH_2_O, and increasing PEEP to 15 cmH_2_O (from a baseline setting of 10 cmH_2_O), while keeping tidal volume fixed at 7 ml/kg, for both the Type 1 CARDS and conventional ARDS models (normal BMI patient).

### Effects of applying ARDSnet PEEP/FiO_2_ tables

To investigate the utility of applying standard combinations of PEEP and FiO_2_ based on tables derived from the ARDSnet trial [27], we evaluated the effect of varying FiO_2_ between 0.4 and 1 at PEEP levels of 5 and 10 cmH_2_O. In the Type 1 CARDS model, at a PEEP of 5 cmH_2_O, application of a FiO_2_ of 0.4 (corresponding to the top of the range suggested in [27]) is insufficient to produce a level of PaO2 exceeding 7.3 kPa; see Figure 2. At PEEP of 10 cmH_2_O, application of FiO_2_ of 0.7 as suggested in [27] is also insufficient to ensure adequate oxygenation in the Type 1 CARDS model. In contrast, application of these settings to the conventional ARDS model produces PaO_2_ levels above the 7.3 kPa threshold for both PEEP values.

**Figure 2:**
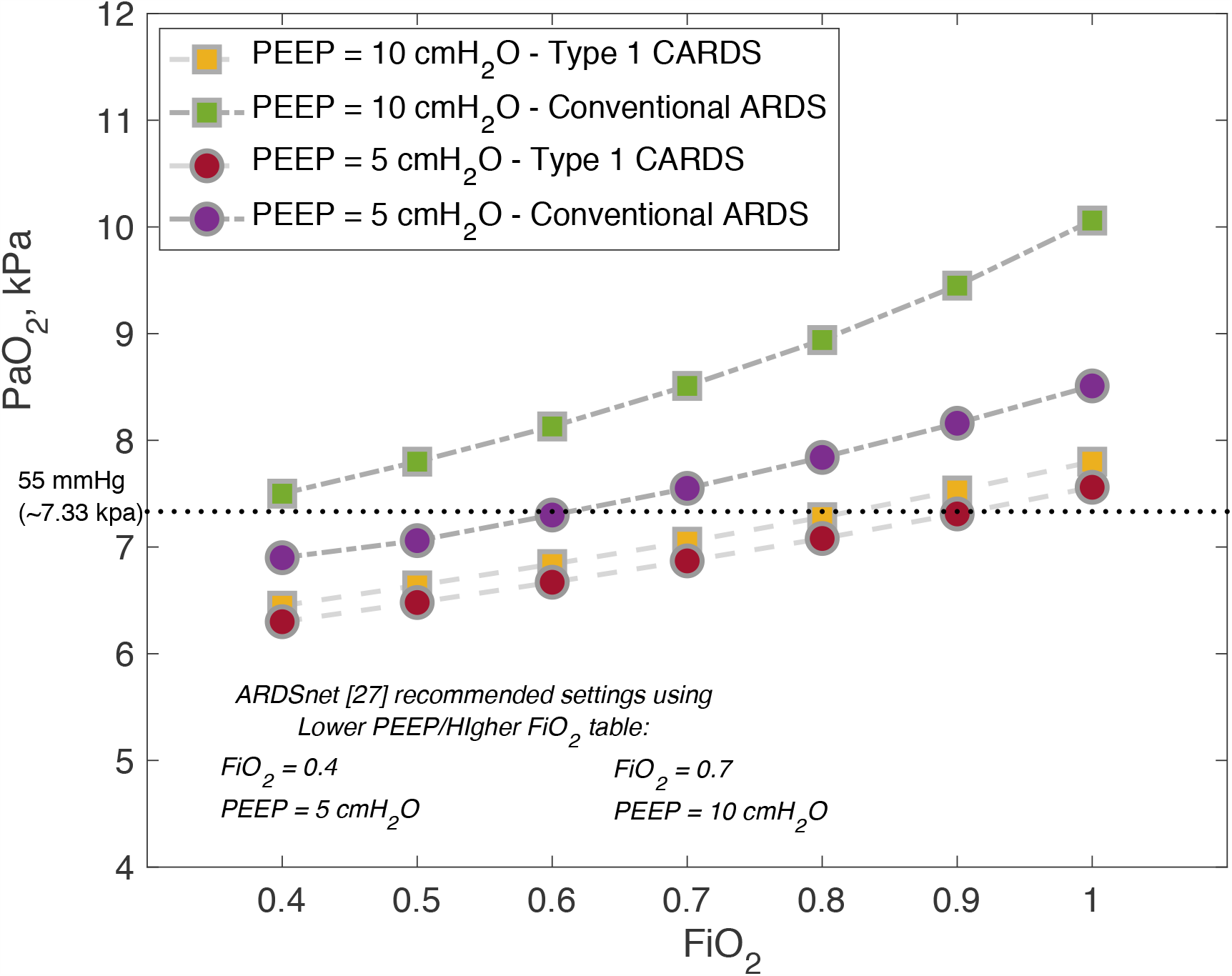
Results of varying FiO_2_ at PEEP levels of 5 and 10 cmH_2_O in both the Type 1 CARDS and conventional ARDS models.

### Effects of high versus low VT

Figure 3 shows the results of reducing VT to 5 ml/kg, and increasing VT to 10 ml/kg (from a baseline setting of 7 ml/kg), while keeping PEEP fixed at 10 cmH_2_O, for both the Type 1 CARDS and conventional ARDS models (normal BMI’s). In this case, increased tidal volume does produce some improvements in gas-exchange in the Type 1 CARDS model, with increased SaO_2_ and DO_2_, and reduced PaCO_2_, although the improvements are smaller than those seen in the conventional ARDS model. Notably, however, the associated worsening in VILI indicators (mechanical power, driving pressure, dynamic strain and plateau pressure) mirrors or exceeds that produced in the conventional ARDS model. Increasing tidal volume in the obese models also produced improved gas-exchange, but again at the cost of clinically significant increases in multiple VILI indices; see Figure S4 in the Supplementary Material. Corresponding results in terms of relative changes, rather than absolute values, for both figures are shown in Figures S5 and S6 of the Supplementary Material.

**Figure 3:**
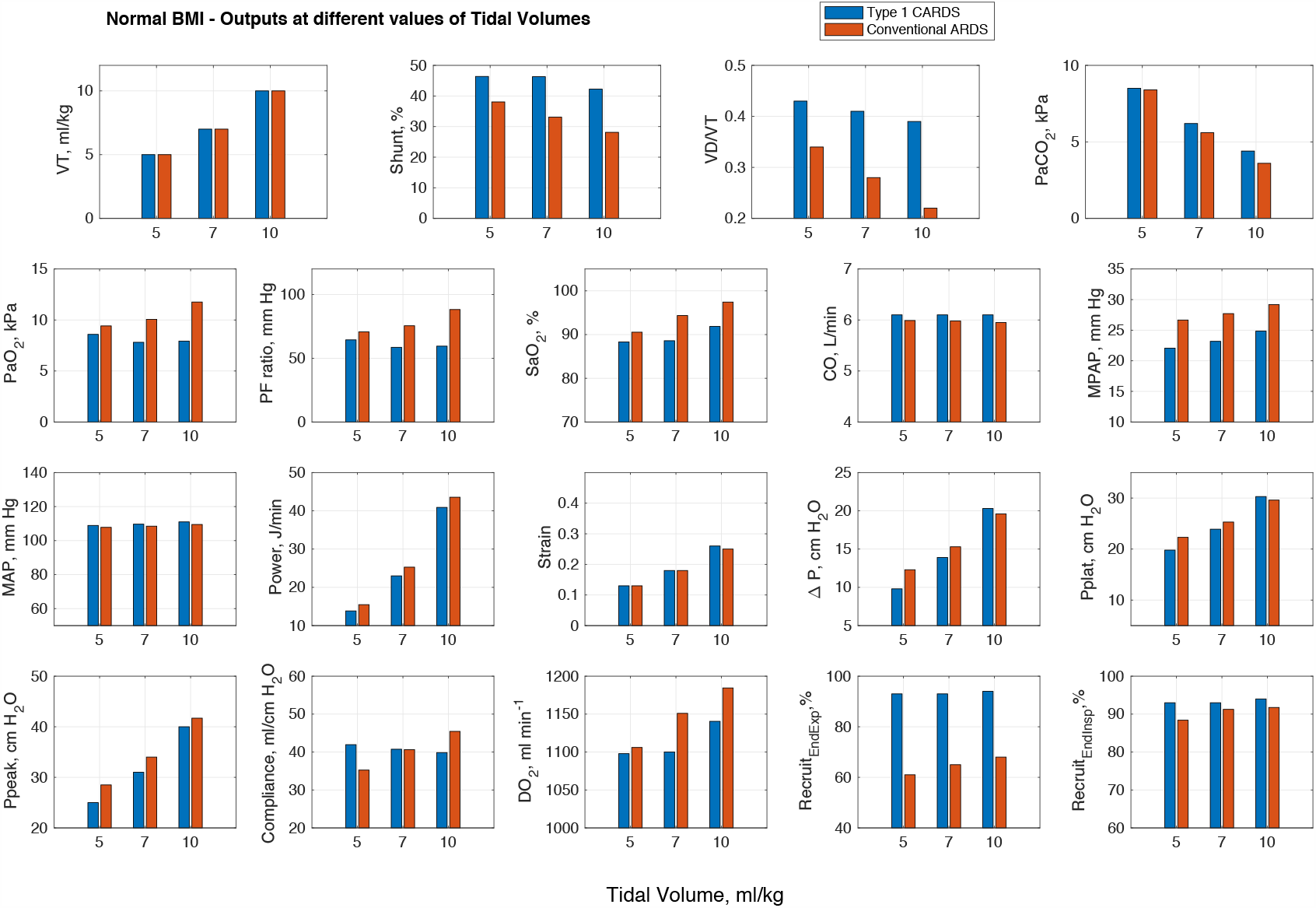
Results of reducing VT to 5 ml/kg, and increasing VT to 10 ml/kg (from a baseline setting of 7 ml/kg), while keeping PEEP fixed at 10 cmH_2_O, for both the Type 1 and conventional ARDS models (normal BMI patient).

## Discussion

The modifications required to our conventional ARDS model to allow it to reproduce the particular features of Type 1 patients are consistent with recent data on lung perfusion abnormalities in CARDS patients from imaging and post-mortem studies. In [16], dual-energy

CT imaging of three CARDS patients revealed preferentially increased perfusion of the lungs proximal to areas of lung opacity, decreased areas of peripheral perfusion corresponding to peripheral lung opacities, and a halo of increased perfusion surrounding peripheral areas of consolidation. A remarkable level of hyperperfusion of gasless tissue was also noted in [1]. A post-mortem study reported vasodilation of pulmonary vessels in two CARDS patients [17]. Finally, two recent studies indicated that CARDS patients are prone to significant vascular dysfunction (also confirmed by the authors’ clinical experience): in [18], 64 clinically relevant thrombotic complications were diagnosed in 150 CARDS patients (including pulmonary embolisms) with >95% of patients having elevated D-dimer and fibrinogen, while a post-mortem examination of 21 CARDS patients in [19] reported massive capillary congestion often accompanied by microthrombi and pulmonary emboli, despite prophylactic anticoagulation.

In our model, we found that implementing pulmonary vasoparesis on its own was not sufficient to produce the levels of shunt and hypoxia, etc. seen in Type 1 patients. However, combining this mechanism with pneumonitis-induced disruption of the alveolar–capillary barrier (as proposed in [20]), and the presence of microthrombi (as reported in [18,19]), allows the model to produce the observed levels of V/Q mismatch and hypoxia, while preserving normal lung compliance and gas volumes. Interestingly, analysis of data from the LUNG SAFE study indicates that a subset of patients with Type 1 characteristics (e.g. poor PEEP responsiveness) also exists in ‘classical’ ARDS; however their frequency is lower than that seen in CARDS [21].

Several studies have examined lung recruitability in small cohorts of CARDS patients using PEEP titration [22-24]. Results reveal a wide variation in the level of recruitability, consistent with our results. Data from these studies suggest that there is potential for PEEP-induced VILI in non-recruitable CARDS lungs (higher driving pressures due to increased PEEP in [23], reduced compliance in [24]), and this is confirmed by the marked elevation in multiple VILI indicators in our Type 1 CARDS model as PEEP is increased from 5 to 15 cm H_2_O. Based on these results, and as recently argued in [25], the use of high PEEP in Type 1 CARDS patients does not appear to be yield a favourable risk/benefit profile; it is therefore essential to evaluate the recruitability of each patient carefully before employing a standard ARDSnet low VT / high PEEP strategy. As demonstrated in Figure 2, application of standard ARDSnet type PEEP/FiO_2_ tables in Type 1 CARDS patients is also unlikely to achieve the levels of oxygenation typically produced in conventional ARDS patients.

It has been asserted that CARDS patients may be able to accept larger tidal volumes than those now normally applied in lung protective ventilation for ARDS without increasing the risk of VILI, [5]. Our results indicate that use of higher VT can produce improvements in oxygenation, but the observed worsening of VILI indicators raises concerns that higher VT could also damage the Type 1 CARDS lung. This may be of particular importance given the relatively longer periods for which CARDS patients appear to require mechanical ventilation [30]. Given the stronger association between improved driving pressure and outcome [26], relative to improved gas-exchange [27], the risks of increasing tidal volume to improve hypoxemia need to be carefully balanced against the increased mortality risk due to higher driving pressure.

The application of prone positioning in patients with CARDS (whether mechanically or non-invasively ventilated) has, in the clinical experience of the authors, a significant beneficial effect on P/F ratios and, albeit anecdotally, on patient outcomes. Globally, intensive care teams are unlikely to have ever simultaneously looked after as many proned patients as they have during the COVID-19 outbreak, and arguably the management of such patients combined with the logistical burden this entailed will be the hallmark of the period. Already of proven benefit in patients with hypoxic respiratory failure [28], prone positioning may have an even more pronounced effect in patients requiring mechanical ventilation due to CARDS. The maneuver has been so successful, at least in the short term, that prone positioning has been extrapolated to spontaneously breathing patients (for the first time in the authors’ experience), resulting in, anecdotally, equally impressive improvements in oxygenation. Prolonged prone positioning has recently been demonstrated to result in improvements in P/F ratios to greater than those recorded pre-intubation in a small study of CARDS patients, with no change in the complication rate [29].

From a physiological standpoint, quite why proning might be so effective in CARDS is not currently understood, particularly in the Type 1 phenotype, where its use remains controversial [1-5]. The argument that proning improves V/Q mismatch through increasing blood flow to functioning ventral alveoli seems to fall short of a full explanation for the significant improvements seen in oxygenation in CARDS. Whilst dual-energy CT scanning of such patients is technically feasible, it is logistically difficult. Use of a physiological simulator such as that developed here may allow us to understand the mechanistic basis for the improvements achieved by proning, and potentially guide the timing, duration and optimization of ventilation for prone positioned CARDS patients.

## Conclusions

A model implementing (a) vasodilation leading to hyperperfusion of collapsed lung regions, (b) disruption of hypoxic pulmonary vasoconstriction, (c) disruption of alveolar gas-exchange due to the effects of pneumonitis, and (d) heightened vascular resistance due to the presence of microthrombi, replicates levels of ventilation-perfusion mismatch and hypoxemia exhibited by Type 1 CARDS patients, while preserving close to normal lung compliance and gas volumes. Our results suggest that ventilatory management of such patients (whether with normal or elevated BMI) should not follow standard protocols, but rather focus on keeping PEEP, and where possible tidal volume, low, in order to avoid the risk of entering the VILI vortex.

## Data Availability

All data referred to in the manuscript is freely available. Please email the corresponding author Professor Declan G. Bates for details.

## Supplementary Materials

1) Supplementary.pdf – Full description of computational simulator, additional figures for elevated BMI patients, additional figures showing relative rather than absolute changes.

## Materials and Correspondence

Requests to be forwarded to Prof. Declan G Bates.

